# Transmission roles of symptomatic and asymptomatic COVID-19 cases: a modeling study

**DOI:** 10.1101/2021.05.11.21257060

**Authors:** Jianbin Tan, Yang Ge, Leonardo Martinez, Jimin Sun, Changwei Li, Adrianna Westbrook, Enfu Chen, Jinren Pan, Yang Li, Feng Ling, Zhiping Chen, Hui Huang, Ye Shen

## Abstract

**Key Points:** *Question:* What is the transmissibility of COVID-19 asymptomatic and symptomatic cases, respectively? To date, they have not been well quantified in existing literature.

*Findings:* The transmissibility of asymptomatic and symptomatic COVID-19 increases with patient age. The asymptomatic cases had a 66.72% lower transmissibility rate than symptomatic cases.

*Meaning:* The transmissibility of asymptomatic COVID-19 cases is not low. Asymptomatic cases are harder to detect compared to symptomatic cases. Consequently, the burden of asymptomatic transmission could potentially dominate the spreading in certain scenarios.

## Background

COVID-19, caused by the novel coronavirus (SARS-CoV-2),[1] is a great threat to human health.[2] Nonpharmaceutical interventions such as social distancing, case isolation and contact quarantines are the most common tools for suppressing the pandemic in many countries where vaccine supplements are limited.[3–5] However, much remains unclear regarding SARS-CoV-2 transmissibility which undermined the efforts for tailored interventions.[6,7] COVID-19 patients may present and remain pre-symptomatic, asymptomatic, or symptomatic and transmission may occur at each of these disease states.[8–10] Unlike the transmission caused by symptomatic cases, pre-symptomatic and asymptomatic transmission are hard to detect and difficult to measure as many surveillance systems rely on symptom-based population screening.[8,11–13] Previous case studies suggested that asymptomatic COVID-19 individuals are less infectious than symptomatic cases.[14,15] However, asymptomatic cases may spread for a longer period due to reduced efficiency in case detection.[16] Several studies investigated the silent transmission of SARS-CoV-2, but presented contradictory conclusions with estimated burden ranged from 3% to 79%.[16–18]

Without sufficient follow-up time, asymptomatic and pre-symptomatic cases are often indistinguishable. Consequently, studies using population-level data to estimate of age-specific transmission and susceptibility parameters commonly falls short of accuracy which potentially explains for the heterogeneous findings from different studies.[19–21] Common issues were modeling without data on observed asymptomatic infection[16–18,22] and inclusion of pre-symptomatic cases as part of an asymptomatic classification,[19–21]. Meanwhile, few studies assessing asymptomatic infectiousness and viral load with limited sample sizes fail to capture the transmission dynamics.[14,15,23–30] No studies to date have attempted to combine reliable case symptom classification with age-dependent transmissibility, social contact measures, and susceptibility parameters at the population level to learn the transmission dynamics. However, a comprehensive understanding of the age-specific symptomatic and asymptomatic transmission dynamics at the population level is essential to the evaluation of an epidemic and the creation of responding health policies.

In this study, we report on a longitudinal cohort of all diagnosed COVID-19 infections, between January 8th and February 23nd, 2020, from Zhejiang province, China. All patients without initial symptoms were followed by at least 90 days to distinguish between asymptomatic and pre-symptomatic cases, an essential procedure rarely implemented by previous studies to ensure reliable classification of case symptoms. We then built age-stratified compartmental models to study the age-dependent population-level transmission roles of symptomatic and asymptomatic COVID-19 cases.

## Methods

### Data sources

Zhejiang province is an eastern coastal province adjacent to Shanghai city with a population of approximately 54 million individuals.[31] The first and only major wave of the COVID-19 epidemic in Zhejiang began on early January, 2020 and continued until late February, 2020 after which only sporadic single-case events were observed. We included information from all confirmed cases in this major wave (a total of 1342 cases), as well as a follow-up investigation related to all detected asymptomatic infections to distinguish between asymptomatic and pre-symptomatic cases. Individual-level data related to the symptom onset of symptomatic infections, as well as COVID-19 confirmation dates and ages of both symptomatic and asymptomatic cases were collected. On January 23rd, 2020, the provincial government changed its infectious disease alert category to the highest level and, on February 1st, began a comprehensive set of interventions.[32] As of April 10th, 2020, the date in which we restricted our data for this analysis, no additional outbreak had been observed. Trained health professionals investigated each confirmed case with a predefined questionnaire by which basic health and demographic information were collected.

### Definition of symptomatic and asymptomatic cases

All confirmed cases and their close contacts were isolated or quarantined after being identified through contact tracing. During the isolation/quarantine period, cases and their contacts received regular testing and daily symptom screening for fever, cough, and shortness of breath. Tests for case confirmation were conducted using reverse transcription polymerase chain reaction (RT-PCR) or viral genome sequencing on samples from throat swabs (oropharynx and nasopharynx). If a case or contact had a positive test result but without any symptoms, they would be temporarily classified as an asymptomatic/pre-symptomatic case at the time. All cases were followed for at least 90 days after their initial positive test to distinguish between asymptomatic and pre-symptomatic cases. Among these subjects, those who developed symptoms later would receive a final classification as a symptomatic case. Others who had never developed any symptoms between their initial positive test and first subsequent negative PCR test would be classified as asymptomatic cases.

### Model structure

We divided the total population of Zhejiang province into seven age groups (Figure 1). To consider transmission related to symptomatic and asymptomatic infections among different age groups, our model contained 8 compartments for *i*^*th*^ the age group: susceptible population (*S*^*i*^), exposed contacts (*E*^*i*^), pre-symptomatic cases (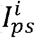, infected but have not yet developed symptoms), symptomatic cases 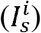, asymptomatic cases (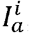, infected but asymptomatic till confirmed/recovery), and removed/recovery groups 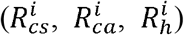. We assumed new infections were driven by transmission from compartments of 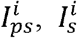 and 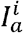 in all age groups.

**Figure 1.**
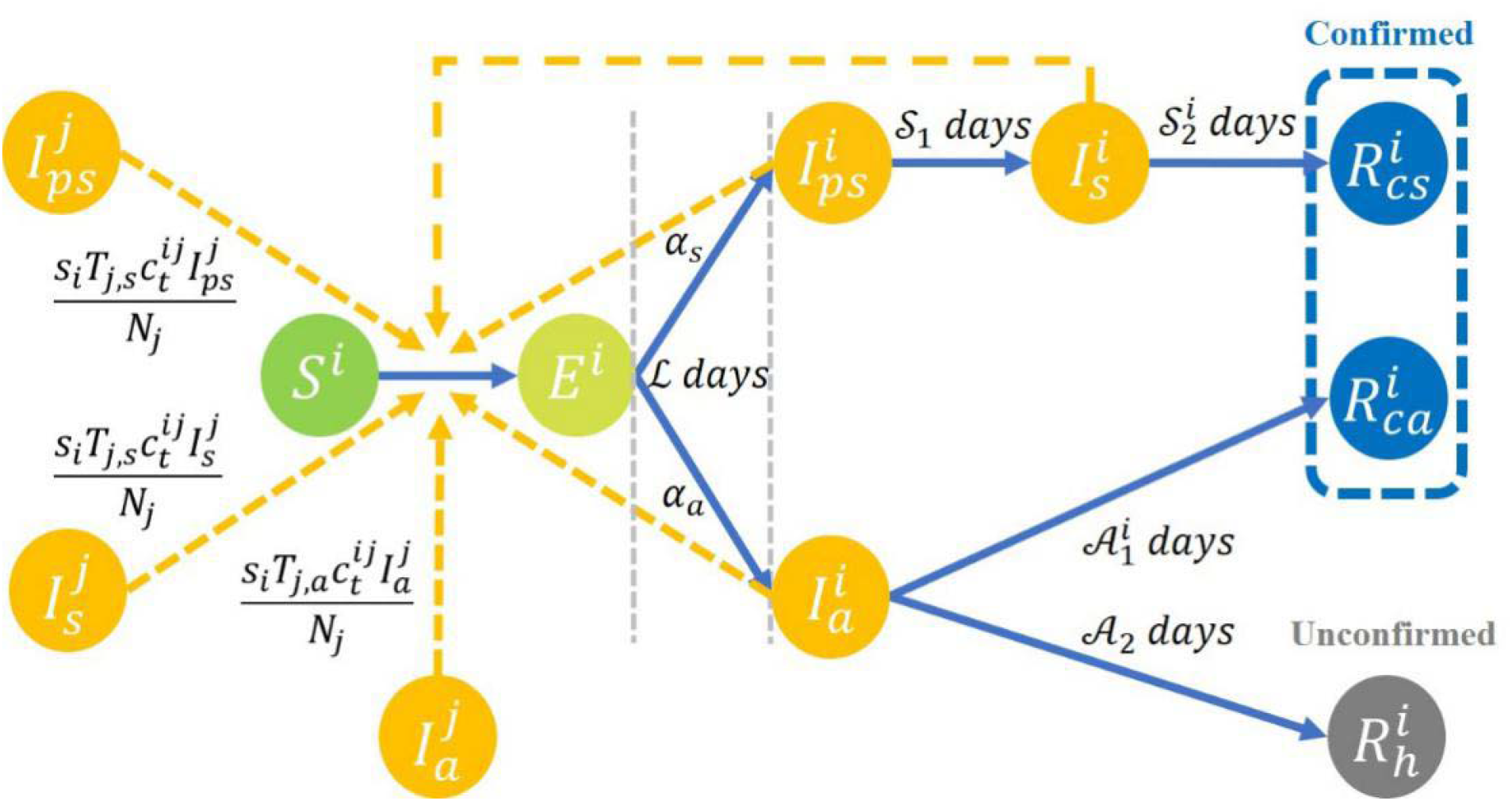
Compartmental model for SARS-CoV-2 transmission, where “*j*” represents another age group different from “*i*” for the compartments.

Asymptomatic cases 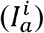 were infections without typical symptoms which were often untraceable in the clinical survey and, therefore, their contribution to population-level transmission would be underestimated. To account for this, we assumed only a proportion of asymptomatic infections were detected 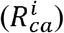, while others 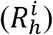 would be unconfirmed. We were able to observe disease confirmation date but not the date of infection for the period from cases becoming infectious to the diagnosis of COVID-19 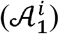 in those with confirmed asymptomatic infection. Based on the virus shedding pattern of asymptomatic infections reported in previous studies,[13,15,33,34] we assumed that this period should be less than 30 days, after which virus shedding generally ceases, and infection is no longer detectable through pathogen-specific testing.

To identify age-varying transmissibility and susceptibility,[22] we assumed a time-varying curve for the average contact numbers of *i*^*th*^ age group with *j*^th^ age group 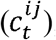, which is estimated with the contact matrix between age groups through surveys conducted in Shanghai.[35,36] We separate the probability of infection into two components: transmissibility (T) and susceptibility (s). We define transmissibility (T) as the infectiousness of one case. Similarly, we define susceptibility (s) as the probability of acquiring infection from an infectious case (T= 1). Therefore, s= 0 corresponds to a situation in which the susceptible individuals are immune to the disease. We assumed that case transmissibility would depend on age and the presence of symptoms. To capture the age-dependent pattern, B splines basis functions were used to model the variability in age-varying transmissibility smoothly. Finally, the compartmental model was fitted to the daily new symptomatic and asymptomatic cases in Zhejiang province for each age group with Markov Chain Monte Carlo (MCMC) algorithm.

All analyses were implemented in R version 3.5.1. Packages of deSolve,[37] extraDistr,[38] and splines[39] were used for model fitting. Unless stated otherwise, the medians of the posterior distributions were used as the point estimators of parameters and simulated numbers.

#### Ethics approval

The research protocol was approved by the institutional review board at the Zhejiang Provincial Center for Disease Control and Prevention. The study was based on deidentified data.

## Results

### Transmissibility

The estimated transmissibility presented an age-dependent difference between symptomatic and asymptomatic infections (Figure 2). While the transmission of symptomatic cases monotonically increased with increasing age, the transmissibility of asymptomatic infection remained low until age 40, after which point it significantly increased with increasing age. The age-varying ratios of the two kinds of transmissibility indicated asymptomatic cases were, on average, 66.72% lower in transmission than symptomatic cases. However, the difference between the two types of infections was not as big in those aged 0-20 and 60+ years old, but became more obvious in the middle-aged group where the ratios were as low as 24.42% and 23.38% for those aged 30-40 and 40-50 years old, respectively.

**Figure 2.**
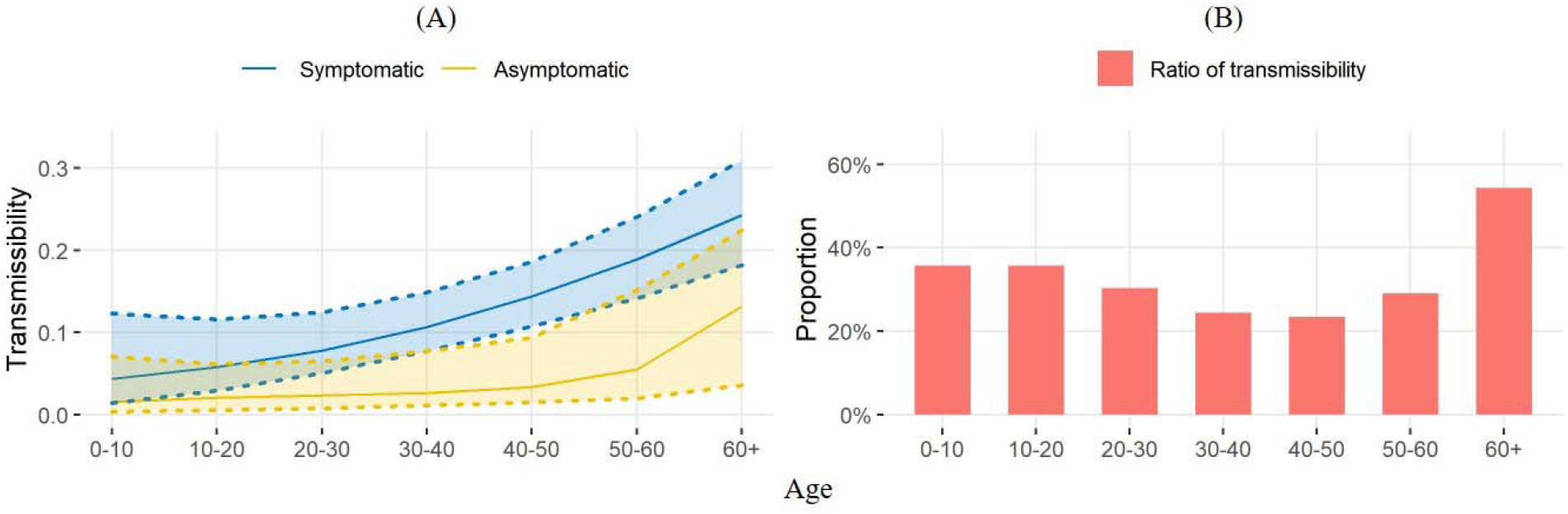
(A) The estimated transmissibility and 95% credible intervals for each age group; (B) The ratios of asymptomatic transmissibility to symptomatic transmissibility for seven age groups.

### The proportion of asymptomatic cases

In Figure 3, the proportion of asymptomatic cases 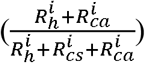 estimated by our model was much larger than what was observed in the data. The average proportion of asymptomatic cases was 28.22% (95%CI: 22.97% - 34.56%) of the total counts of cases in our model estimation, but was 9.24% in the observed data 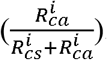. In our estimation from the empirical data, the highest proportion of asymptomatic case was among 0-10 (60.18% (95%CI: 53.61% - 66.99%)) and 10-20 (57.64% (95%CI: 47.45% – 66.98%)) years old groups. For asymptomatic cases, we further estimated the proportion of cases that failed to be detected 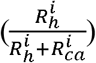. In the posterior samples, the average proportion of unconfirmed cases in all asymptomatic infections was 74.10% (95%CI: 65.85% - 80.72%). The maximum proportion of unconfirmed cases was observed in 20-30 years old at 86.59% (95%CI: 73.64% - 92.19%).

**Figure 3.**
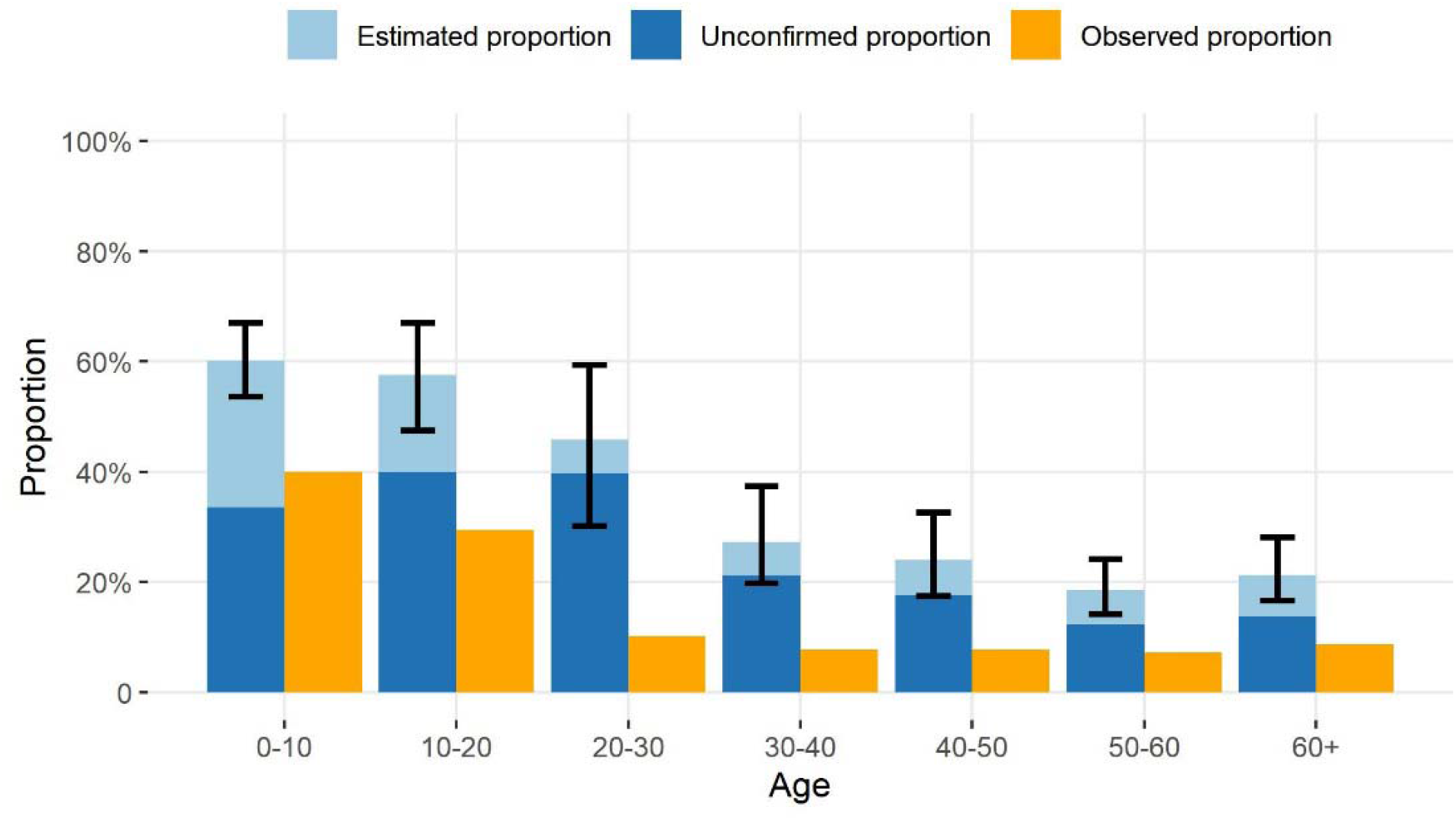
The proportion of asymptomatic infections and unconfirmed asymptomatic infections until February 22nd, 2020, for seven age groups. The estimated proportions of asymptomatic cases, the proportions of cases that failed to be detected among asymptomatic infections (unconfirmed proportions), and the observed proportions of asymptomatic cases are defined as: 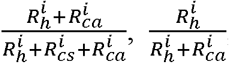, and 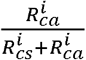 respectively. The 95% credible intervals for the estimated proportions of asymptomatic cases are shown for each age group.

### Symptomatic and asymptomatic transmission

To explore the impact of symptomatic and asymptomatic transmission, we present several features of the estimated dynamic of the epidemic and the transmission burden caused by symptomatic and asymptomatic cases in Figure 4. The estimated number of daily new transmissions reached a peak around ten days prior to the peak of the daily reported new confirmed cases (Figure 4A). We estimated a substantial number of undetected asymptomatic cases (109 (95%CI: 73 - 164)) were infected before the first asymptomatic case was diagnosed (January 27th) (Figure 4B). New transmissions were nearly eliminated by February 2nd, 2020 (Figure 4A), when a comprehensive set of restrictions had been implemented. The peak of the two types of transmission both occurred between January 18th to 22nd (Figure 4C). The average burden of asymptomatic transmission during the major outbreak period was estimated to be 12.86% (95%CI: 7.54% - 19.27%). The burden of asymptomatic transmission increased with time, ranging from 7.77% to 16.03% (Figure 4D). Simulation studies were conducted to investigate the dynamic changes in the transmission burden over time during a prolonged epidemic. When the duration of the decreasing process of the contact function (represented by “*m*”) was prolonged by two weeks and each individual’s daily contact number was increased by one person during the outbreak period, we found a slower decreasing trend in daily new cases infected by asymptomatic cases compared with that contributed by symptomatic cases. Additional scenarios were generated demonstrating the possibility of asymptomatic transmission dominating the total transmission under different conditions, especially when the duration between symptom onset and disease confirmation for symptomatic infections was shortened and the asymptomatic infections were not controlled.

**Figure 4.**
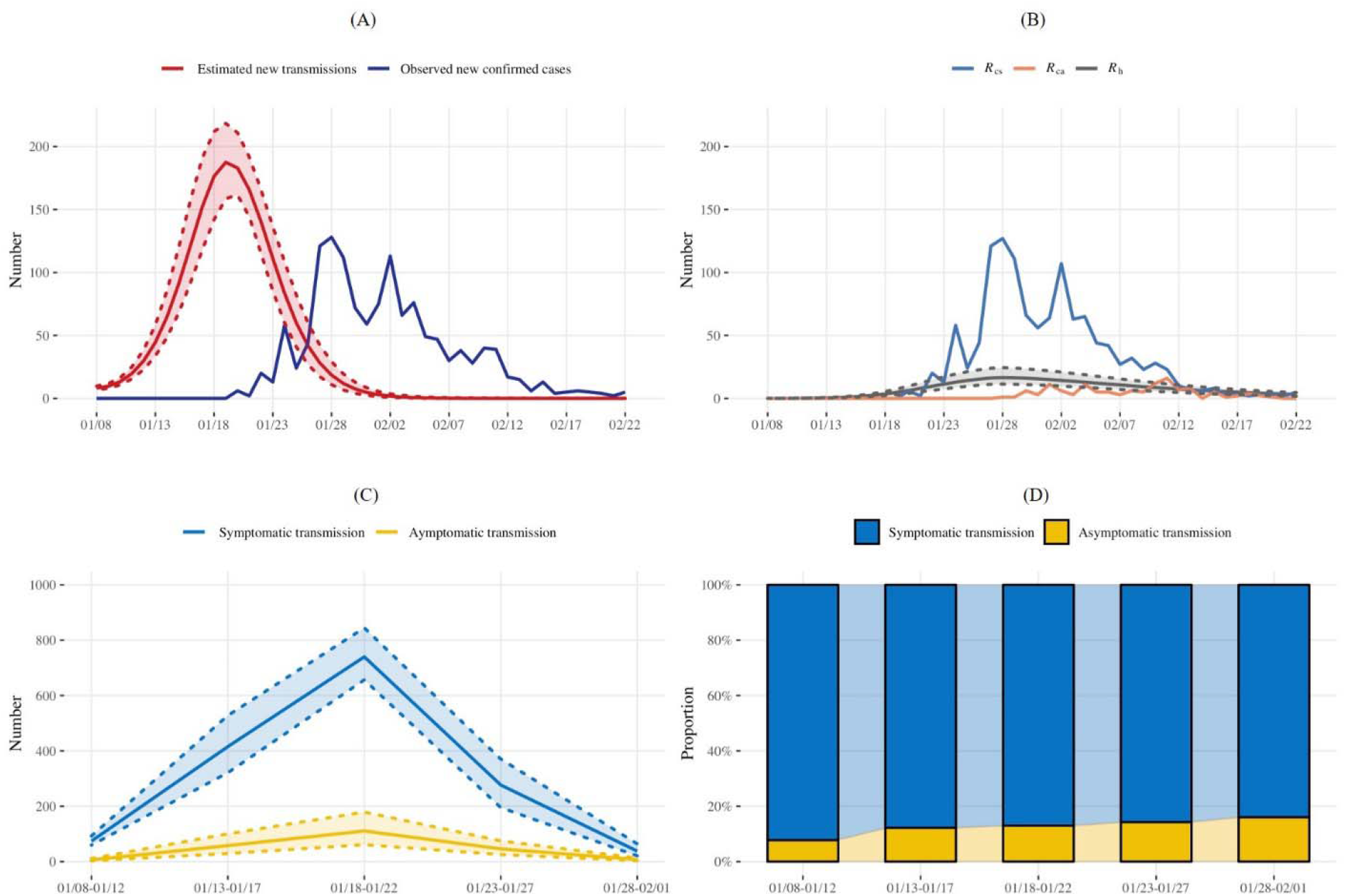
The estimated dynamics of the epidemic and the transmission burdens from symptomatic and asymptomatic cases. (A): The estimated numbers of daily new transmissions with 95% credible intervals and the observed numbers of daily reported new confirmed cases from January 8th to February 22nd, 2020; (B) The observed numbers of daily reported new confirmed symptomatic (*R*_*cs*_) and asymptomatic cases (*R*_*ca*_) and the estimated numbers of daily new cases that failed to be detected (*R*_*h*_) with 95% credible intervals; (C): The estimated numbers of infected individuals caused by symptomatic and asymptomatic transmission over time, with 95% credible intervals; (D): Corresponding proportions of symptomatic and asymptomatic transmissions over different time periods.

### Age-depended transmission

Within each age group, we observed heterogeneous transmission contributions during different time periods (Figure 5A). Early on in the epidemic, the transmission burden was dominated by persons of 50-60 years old (32.75% from January 8th to January 12th), but the proportion of transmission contribution from people over 60 years old significantly increased over time, surpassing the 50-60 years old and reaching 30.42% by February 1st, 2020. The proportion of transmission contribution among varying age groups was distinct between symptomatic and asymptomatic cases (Figure 5B). The majority of both symptomatic and asymptomatic transmissions were contributed by persons over 30 years old (Table S9). Individuals below 30 years old only contributed less than 5% of all symptomatic transmission and approximately 12% of all asymptomatic transmission, respectively, despite representing almost 40% of the entire population. Contributions to asymptomatic transmission among 20-30 and >60 year age groups (9.44%, and 31.73%, respectively) were substantially higher than their corresponding contributions to symptomatic transmission (3.77%, and 26.55%, respectively). To further understand possible age-dependent vaccination strategies, a simulation of seven scenarios was conducted to assess the percentage decline in different age groups if one age group were to achieve 100% immunity by vaccinations. The results suggested that vaccinations targeting age groups above 30 years are likely to be more effective at the population level, with the most percentage decline of cases from the entire population achieved by targeting the 50-60 years old group. Meanwhile, vaccinating those younger than 30 years old are more likely to benefit their own age groups.

**Figure 5.**
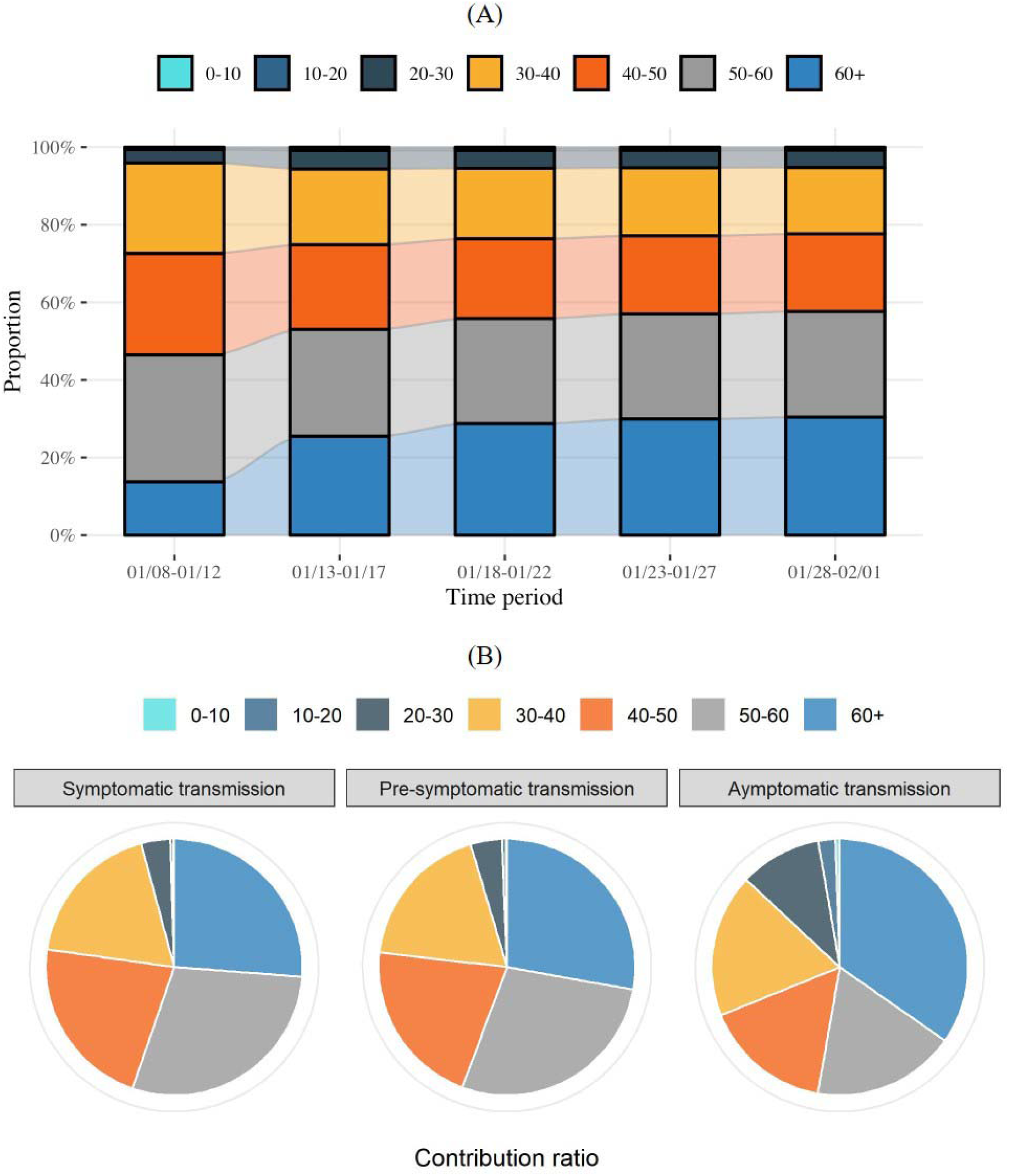
The burden of transmission caused by different ages. (A) The estimated (contribution) ratios of new transmissions from different ages over different time periods; (B) The estimated (contribution) ratios of symptomatic and asymptomatic transmission from different ages. The contribution ratio of each age group is calculated by the proportion of the transmissions caused by the corresponding age group to the number of all transmissions in each transmission type, from January 8th to February 1st, 2020.

## Discussion

In our study, we found that asymptomatic cases were over 60% less infectious compared to symptomatic cases. While great efforts like mass screening and strict contact tracing were conducted, our results suggested that a large proportion of asymptomatic infections were still not detected.[40] The burden of asymptomatic transmission was inferior in the early outbreak but could become higher with the continuous spread of COVID-19. Under strained resources, age-specific prevention and control strategies aimed at the middle age population may return greater population-level benefits.

Current evidence suggests that asymptomatic COVID-19 cases are generally less infectious[14] than cases with symptoms. We found that this difference may partially be explained by patient age. Age may directly impact COVID-19 transmission through virus shedding patterns[15] as discussed in previous studies.[41] Symptoms are commonly mild in children[42] but severe in the elderly.[43] While still debatable,[44] higher severity has been associated with increased shedding of the virus.[45] In our study, symptomatic and asymptomatic cases were most infectious in individuals 60 years old or older. In contrary to the monotonic increasing association between age and transmission in symptomatic cases, there was a plateau of a low degree of transmission in young asymptomatic infections. We suspect older adults are not only the most vulnerable to succumb to COVID-19 but also may be more likely to transmit once infected, regardless of symptom status. Interventions attempting to suppress asymptomatic transmission, such as mask-wearing, should primarily focus on older adults if interventions to the whole community are not feasible.

Similar to previous studies, our results suggest a small proportion of asymptomatic cases have been detected since the start of the COVID-19 pandemic in our setting.[46–48] Symptom-based screening has limited capability in asymptomatic case detection,[6] while mass pathogen or immunological-based testing at the population-level consumes tremendous health resources, and thus is not feasible in most settings. Considering these challenges, age-dependent screening strategies may be more practical. We found that the highest number of undetected asymptomatic cases was among young adults aged 20 to 30 years old (Table S7) and the corresponding transmission contribution was significantly higher than that of symptomatic case (Figure 5B). Young people were less likely to adhere to social distancing guidelines,[49] often had mild symptoms or were asymptomatic after infection,[50] and were not prioritized in prevention and controlling strategies.[51] Meanwhile, younger asymptomatic patients were also more likely to have normal CAT scan findings, which may further complicate case detection.[12] Case detection of asymptomatic COVID-19 cases based on current control strategies implemented in this study is alarming.

Based on the estimated transmission contributions from symptomatic and asymptomatic infections, roughly 13% of infections were associated with asymptomatic transmission and that percentage continuously increased with a prolonged period. The overall burden of transmission was mainly contributed by symptomatic cases at the beginning of the epidemic, but asymptomatic infections appeared to have increasing percentages of subsequent cases later on. Additional simulations suggested that the transmission burden could even be dominated by asymptomatic transmissions under certain circumstances. Therefore, the spreading potential of asymptomatic cases cannot be ignored, especially in the later stages of the epidemic and in regions where social distancing has not been mandated. Meanwhile, potential differences in transmission burden by age groups, as shown in Figure 5 and S6, supports prioritizing age-dependent prevention and control strategies when facing strained resources. As the larger contributor to the transmission of COVID-19, the older age population is not only a highly vulnerable group but should also be the primary target for prevention strategies. Vaccine Strategies prioritizing the population between 30-60 years old are likely to have the most population-level benefits.

There are several limitations in this study. First, data collection likely missed potential cases of the epidemic, despite intensified efforts devoted by the local investigation team to trace contacts. Due to this, we introduced a compartment in our model 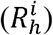 to adjust for poor case ascertainment and missing cases. Second, transmissibility and susceptibility were two factors related to symptomatic and asymptomatic transmission estimation and can be difficult to capture simultaneously. We used the susceptibility estimates from a previous study[22] as priors in our model to account for this parameter identification problem. Third, the contact survey data we used in our model were obtained in Shanghai, a city adjacent to Zhejiang province. Although the two regions share a similar culture and modes of social activities, there were potential uncertainties associated with the discrepancies in contact matrices. To address this limitation, we introduced a correction parameter in our model, so these uncertainties were partially adjusted for in the analyses.

## Conclusion

In summary, our results suggest individual-level transmissibility of COVID-19 increases with patient age, therefore targeting older age groups with prevention and intervention strategies is expected to be more efficient. While asymptomatic cases are difficult to trace, the burden of asymptomatic transmission is still sizable and should not be ignored. The results from our study can be used to inform policy decisions on pandemic control and safe reopening.

## Data Availability

The datasets are available from the corresponding author on reasonable request.

