## Supplementary Appendix for "Transmission roles of symptomatic and asymptomatic COVID-19 cases: a modeling study" (from another version of this preprint: version 1, 2021-05-14)

### Contact function

Although the changing mechanism of social contact between the two cross-sectional surveys was not observed, we assume that it could be described by a function of time (days). We denote the average contact number of the $i^{th}$ with $j^{th}$ age group at time t as $c_{t}^{ij}$, and assume that $c_{t}^{ij}$is a constant at the baseline period of the epidemic and decreases toward the stable level during the outbreak period, as reported cases and the implementations of the nonpharmaceutical interventions increases. In particular, the monotonic decline function $c_{t}^{ij}$ followed the supplied equation^1^:

$c_{t}^{ij}=c_{0}^{ij}\left( \frac{\text{1-}\eta_{ij}}{1+\exp(\lambda_{m}(t-d-\frac{m}{2}))}+\eta_{ij} \right)$	(S1)

where $c_{0}^{ij}$ denotes the maximum contact number at an early stage of the outbreak, $d$ represents the time that $c_{t}^{ij}$ starts declining, $\eta_{ij}$ is the ratio of $c_{0}^{ij}$ to the minimum of $c_{t}^{ij}$, which is $c_{0}^{ij}\eta_{ij}$. We chose $\lambda_{m}$ as $\frac{2\text{log}(99)}{m}$ to ensure $m$ represents the duration of the decreasing process of $c_{t}^{ij}$. An illustrative graph of the contact function is given in Figure S1:

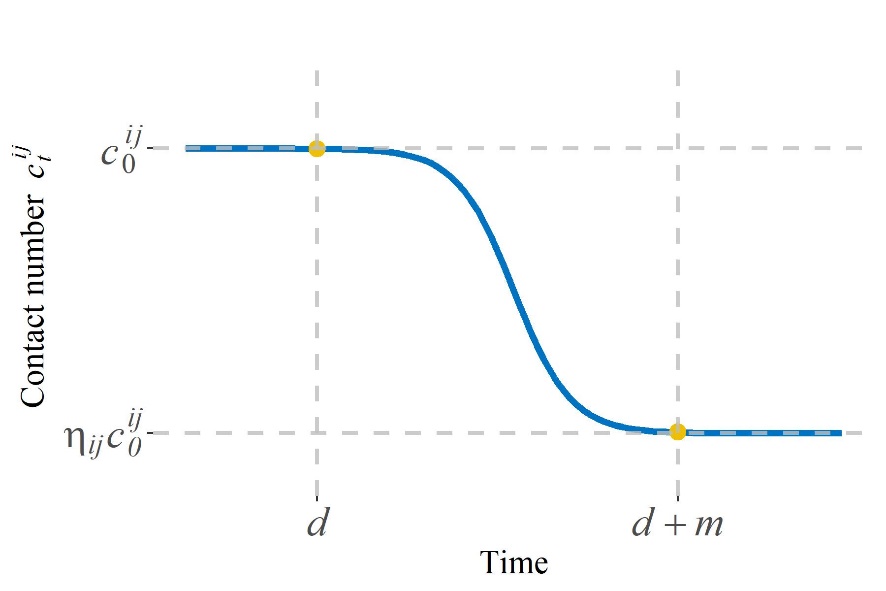

*Figure S1. The graph of contact function*

where $c_{0}^{ij}$ and $\eta_{ij}c_{0}^{ij}$ were the contact numbers of $i^{th}$ with $j^{th}$ age group in the baseline period and outbreak period, respectively.

Notice that the contact matrix of Shanghai during the baseline period and outbreak period^2,3^ was stratified into 5-year age bands, so the two cross-sectional surveys would not be directly used in modeling of the contact function. We firstly proposed a transformation procedure to stratify them into 10-year age bands. To demonstrate this process, take 0-10 years old as an example. Let $c_{b}^{11}$, $c_{b}^{12}$ be the average contact numbers of 0-5 years old with 0-5 years old and 5-10 years old in the baseline period, respectively. Similarly, let $c_{b}^{21}$, $c_{b}^{22}$ be the average contact numbers of 5-10 years old with 0-5 years old and 5-10 years old in the baseline period, respectively. We calculated the average contact numbers of 0-10 years old with 0-10 years old on baseline period ($c_{b,raw}^{11}$) by:

$$\begin{matrix} c_{b,raw}^{11}=\frac{N_{1}}{N_{1}+N_{2}}\cdot(c_{b}^{11}+c_{b}^{12})+\frac{N_{2}}{N_{1}+N_{2}}\cdot(c_{b}^{21}+c_{b}^{22}), \end{matrix}$$

where $N_{1}$, $N_{2}$ is the number of total populations in 0-5 and 5-10 years old in Zhejiang province, respectively.

After the transformation procedure, let $c_{b,raw}^{ij}$ and $c_{o,raw}^{ij}$ be the transformed average contact numbers of $i^{th}$ age group with $j^{th}$ age group in baseline period and outbreak period. To correct the potential discrepancy in Zhejiang province for the use of the contact numbers in the baseline period and outbreak period of Shanghai, we assume that:

$$c_{0}^{ij}=\beta_{1}c_{b,raw}^{ij},$$

$$\eta_{ij}c_{0}^{ij}=\beta_{2}c_{o,raw}^{ij},$$

where $\beta_{1}$ and $\beta_{2}$ are correction parameters for the contact number in baseline period and outbreak period for Zhejiang province. These two parameters can be later estimated when we specify the compartmental model.

#### B-spline approximations for modeling the transmissibility of symptomatic infections

Transmissibility was used to quantify the infectiousness of an infected individual. We assume that the infectiousness is related to the virus shedding ability of a case, which may be associated with age^4^. Therefore, we apply a nonparametric estimation approach using the basis expansion approximation. We use three basis functions, as illustrated in Figure S2, to capture possible shapes of transmissibility for different age groups. Specifically, denoting the transmissibility of the $i^{th}$ ages group as $T_{i,s}$, $\mathrm{it}$ can be approximated as:

$T_{i,s}=\sum_{k=1}^{K} \omega_{k}f_{k}(i)$, $\omega_{k}\geq0$, (S2)

where $f_{k}$ are B-spline basis functions (Figure S2) generated by the quadratic piecewise polynomials (with intercept) valued in $\left[ 0, 8 \right]$ through R package **splines**.

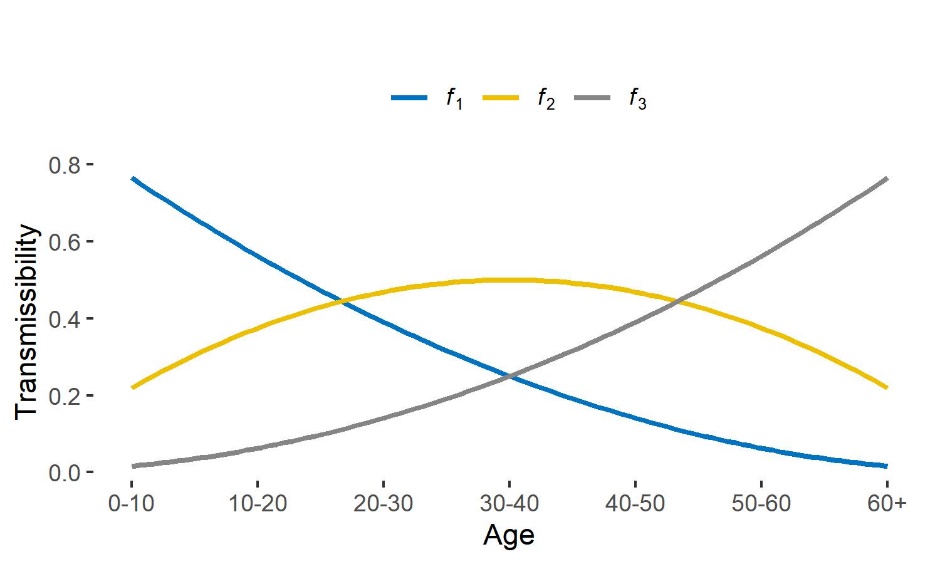

Figure S2. The graph of B-spline.

#### The dynamic system of the compartmental model

Let $S^{i}(t), E^{i}(t), I_{ps}^{i}(t), I_{s}^{i}(t), I_{a}^{i}(t), R_{cs}^{i}(t), R_{ca}^{i}(t), R_{h}^{i}(t)$ be the numbers of corresponding compartments at time $t$ in the $i^{th}$ ages group, the dynamic system of the compartments of different ages is assumed to follow:

$$\begin{matrix} \frac{\text{d}S^{i}\left( t \right)}{\text{d}t} & =-S^{i}\left( t \right)\frac{s_{i}\sum_{j} c_{t}^{ij}\left[ T_{j,s}\left( I_{ps}^{j}\left( t \right)+I_{s}^{j}\left( t \right) \right)+T_{j,a}I_{a}^{j}\left( t \right) \right]}{N_{j}}, \\ \frac{\text{d}E^{i}\left( t \right)}{\text{d}t} & =S^{i}\left( t \right)\frac{s_{i}\sum_{j} c_{t}^{ij}\left[ T_{j,s}\left( I_{ps}^{j}\left( t \right)+I_{s}^{j}\left( t \right) \right)+T_{j,a}I_{a}^{j}\left( t \right) \right]}{N_{j}}-\frac{E^{i}\left( t \right)}{\mathcal{L}}, \\ \frac{\text{d}I_{ps}^{i}\left( t \right)}{\text{d}t} & =\frac{E^{i}\left( t \right)}{\mathcal{L}}\alpha_{s}^{i}-\frac{I_{ps}^{i}\left( t \right)}{\mathcal{S}_{1}},\frac{\text{d}I_{s}^{i}\left( t \right)}{\text{d}t}=\frac{I_{ps}^{i}\left( t \right)}{\mathcal{S}_{1}}-\frac{I_{s}^{i}\left( t \right)}{\mathcal{S}_{2}^{i}}, \\ \frac{\text{d}I_{a}^{i}\left( t \right)}{\text{d}t} & =\frac{E^{i}\left( t \right)}{\mathcal{L}}\alpha_{a}^{i}-\frac{I_{a}^{i}\left( t \right)}{\mathcal{A}_{1}^{i}}-\frac{I_{a}^{i}\left( t \right)}{\mathcal{A}_{2}}, \\ \frac{\text{d}R_{cs}^{i}\left( t \right)}{\text{d}t} & =\frac{I_{s}^{i}\left( t \right)}{\mathcal{S}_{2}^{i}}, \\ \frac{\text{d}R_{ca}^{i}\left( t \right)}{\text{d}t} & =\frac{I_{a}^{i}\left( t \right)}{\mathcal{A}_{1}^{i}}, \\ \frac{\text{d}R_{h}^{i}\left( t \right)}{\text{d}t} & =\frac{I_{a}^{i}\left( t \right)}{\mathcal{A}_{2}}, \end{matrix} (S3)$$

where $\alpha_{s}^{i}+\alpha_{a}^{i}=1$. Note that $N_{i}$ and $s_{i}$ are the total population and susceptibility of the $i^{th}$ age group, respectively, and $c_{t}^{ij}$ is the average contact numbers of $i^{th}$ age group with $j^{th}$ age group at time $t$, which is determined by equation (S1) given $c_{b,raw}^{ij}$, $c_{o,raw}^{ij}$, *d, m,* $\beta_{1}$ and $\beta_{2}$. In addition, $T_{i,s}$ is the transmissibility of symptomatic infected individuals in the $i^{th}$ age group specified by equation (S2) given the parameters $\omega_{1}, \omega_{2}, \omega_{3}$. We further use $T_{i,a}=\gamma_{i}T_{i,s}$ as the transmissibility of asymptomatic individuals in $i^{th}$ age group. Moreover, $\mathcal{L,}\mathcal{S}_{1}, \mathcal{A}_{2}$are the average lengths of the latent period, the pre-symptomatic period, and the period from the beginning of asymptomatic transmission to self-recovery for an asymptomatic infection, respectively. Likewise, $\mathcal{S}_{2}^{i}$ represents the mean duration of the period of symptomatic transmission and $\mathcal{A}_{1}^{i}$ represents the period between the beginning of asymptomatic transmission and the diagnosis of asymptomatic infection in the $i^{th}$ age group.

Based on the equation system (S3), we can calculate the infected numbers caused by symptomatic and asymptomatic transmission for different ages, which is achieved by defining new compartments:

$$\frac{dE_{s}^{j}\left( t \right)}{dt}=\sum_{i} S^{i}\left( t \right)\frac{s_{i}T_{j,s}c_{t}^{ij}\left( I_{ps}^{j}\left( t \right)+I_{s}^{j}\left( t \right) \right)}{N_{j}}, \left( S4 \right)$$

$$\frac{dE_{a}^{j}\left( t \right)}{dt}=\sum_{i} S^{i}\left( t \right)\frac{s_{i}T_{j,a}c_{t}^{ij}I_{a}^{j}\left( t \right)}{N_{j}}, (S5)$$

where $E_{s}^{j}\left( t \right)$ and $E_{a}^{j}\left( t \right)$ represent the number of the infected individuals caused by symptomatic and asymptomatic transmission from $j^{th}$ age group, respectively. Similarly, we define $E_{s}\left( t \right)=\sum_{j} E_{s}^{j}\left( t \right)$ and $E_{a}\left( t \right)=\sum_{j} E_{a}^{j}\left( t \right)$ as the number of infected individuals caused by symptomatic and asymptomatic transmissions. We also use the ratios $\frac{E_{a}}{E_{s}+E_{a}}, \frac{E_{s}^{j}}{E_{s}}$ and $\frac{E_{a}^{j}}{E_{a}}$to quantify the burden of asymptomatic transmission, the contribution of $j^{th}$age group among symptomatic and asymptomatic transmissions, respectively.

#### Bayesian modeling

A Bayesian paradigm provides a coherent probabilistic approach to introduce reasonable uncertainties into the modeling of epidemic process^5^. In particular, let

$$\begin{matrix} Z(t) & :=(S^{i}(t),E^{i}(t),I_{ps}^{i}(t),I_{s}^{i}(t),I_{a}^{i}(t),R_{cs}^{i}(t),R_{ca}^{i}(t),R_{h}^{i}(t))_{i=1:7}, \\ \Omega: & =(\mathcal{L},\mathcal{S}_{1},\mathcal{A}_{2},d,m,\beta_{1},\beta_{2},\omega_{1},\omega_{2},\omega_{3},(s_{i},\alpha_{a}^{i},\mathcal{S}_{2}^{i},\mathcal{A}_{1}^{i},\gamma_{i})_{i=1:7}), \end{matrix}$$

where $\Omega$ is the collection of all parameters needed to be estimated. As mentioned above, we assume that$\mathcal{C}_{b,raw}:=(c_{b,raw}^{ij})_{i,j=1:7}$, $\mathcal{C}_{o,raw}:=(c_{o,raw}^{ij})_{i,j=1:7}$, the estimated means of the age-varying susceptibility ${(s_{i,\mu})}_{i=1:7}$and $N:=(N_{i})_{i=1:7}$ are known, which are shown in Table S1-S4:

Table S1. Transformed contact matrix (10-year age bands) during the baseline period ($\mathcal{C}_{b,raw})$, consisting of the average number of contacts per day.

| Age | | Age of contact | | | | | | |
| --- | --- | --- | --- | --- | --- | --- | --- | --- |
|  |  | 0-10 | 10-20 | 20-30 | 30-40 | 40-50 | 50-60 | 60+ |
| Age of participant | 0-10 | 8.31 | 0.93 | 0.69 | 2.11 | 0.35 | 0.57 | 1.34 |
|  | 10-20 | 0.48 | 19.17 | 1.35 | 3.13 | 2.51 | 1.14 | 1.06 |
|  | 20-30 | 0.16 | 0.21 | 4.38 | 5.66 | 5.33 | 3.28 | 1.77 |
|  | 30-40 | 0.60 | 0.45 | 3.14 | 6.88 | 5.28 | 3.20 | 1.74 |
|  | 40-50 | 0.12 | 0.68 | 2.80 | 5.44 | 5.34 | 2.69 | 1.99 |
|  | 50-60 | 0.17 | 0.24 | 2.34 | 3.91 | 3.91 | 3.80 | 3.96 |
|  | 60+ | 0.13 | 0.13 | 0.59 | 1.22 | 1.53 | 1.90 | 6.12 |

Table S2. Transformed contact matrix (10-year age bands) during the outbreak period $(\mathcal{C}_{o,raw})$, consisting of the average number of contacts per day.

| Age | | Age of contact | | | | | | |
| --- | --- | --- | --- | --- | --- | --- | --- | --- |
|  |  | 0-10 | 10-20 | 20-30 | 30-40 | 40-50 | 50-60 | 60+ |
| Age of participant | 0-10 | 0.02 | 0.00 | 0.28 | 1.41 | 0.16 | 0.07 | 0.00 |
|  | 10-20 | 0.11 | 0.22 | 0.48 | 0.53 | 1.10 | 0.43 | 0.37 |
|  | 20-30 | 0.07 | 0.07 | 0.24 | 0.16 | 0.37 | 0.65 | 0.21 |
|  | 30-40 | 0.40 | 0.17 | 0.17 | 0.71 | 0.35 | 0.16 | 0.52 |
|  | 40-50 | 0.21 | 0.56 | 0.26 | 0.33 | 0.77 | 0.34 | 0.51 |
|  | 50-60 | 0.06 | 0.06 | 0.42 | 0.17 | 0.20 | 0.52 | 0.42 |
|  | 60+ | 0.05 | 0.06 | 0.08 | 0.24 | 0.08 | 0.07 | 0.83 |

Table S3. The estimated mean of the age-varying susceptibility^22^ ${(s_{i,\mu})}_{i=1:7}$

| Age | 0-10 | 10-20 | 20-30 | 30-40 | 40-50 | 50-60 | 60+ |
| --- | --- | --- | --- | --- | --- | --- | --- |
| $s_{i,\mu}$ | 0.40 | 0.38 | 0.79 | 0.86 | 0.80 | 0.82 | 0.81^[1]^ |

Table S4. The age structure of Zhejiang province^6^ $(N)$

| Age | 0-10 | 10-20 | 20-30 | 30-40 | 40-50 | 50-60 | 60+ |
| --- | --- | --- | --- | --- | --- | --- | --- |
| Population | 4811720 | 5969042 | 9417394 | 9632456 | 10201561 | 6836085 | 7558633 |

Let $D_{t}^{i,1}$ be the number of daily new symptomatic infected individuals, as defined by the onset of symptoms, at time $t$ in $i^{th}$ age group, and $D_{t}^{i,2}$ be the number of daily new confirmed asymptomatic infected individuals at time $t$ in $i^{th}$ age group. Similarly, $\mathcal{D}_{t}^{i,g}(Z(0),\Omega) (g=1,2$) represents the corresponding fitted values of the epidemic equation (S3) given through the initial value $Z(0)$ and the parameters $\Omega$, where $Z(0)$ is specified by the initial cases of symptom onset on January 7^th^, 2020 and population numbers in Table S4. For a flexible adjustment of overdispersion of the new numbers, we use a negative binomial mass as the loss function for $D_{t}^{i,g}$:

$\begin{matrix} \text{Lik}=\Pi_{t=1}^{T}\Pi_{i=1}^{7}\Pi_{g=1,2}\text{Nb}(D_{t}^{i,g}|\mathcal{D}_{t}^{i,g}(Z(0),\Omega),v_{g}), \end{matrix}$

Where Lik means the likelihood of data, $T$ is the time length of the observed data and $\text{Nb}(u,v)$ represents the mass function of the negative binomial distribution with mean $u$ and size $v$. The note of these parameters is given in Table S5.

Table S5. The note of parameters

| Parameters | Meaning | Reference value | Prior distribution |
| --- | --- | --- | --- |
| $\mathcal{L}$ | Latent period | 2 days^7,8^ | $\log N(2,\sigma)$ |
| $\mathcal{S}_{1}$ | The period of pre-symptomatic transmission | 3-4 days^9,10^ | $\log N(3,\sigma)$ |
| $\mathcal{S}_{2}^{i}$ | The period of symptomatic transmission | Null | $IG(a_{i}, b_{i})$ |
| $\mathcal{A}_{1}^{i}$ | The period from the beginning of asymptomatic transmission to diagnosis for an asymptomatic infection | Null | $Unif(\frac{b_{i}}{a_{i}}, 30)$ |
| $\mathcal{A}_{2}$ | The period from the beginning of asymptomatic transmission to recovered for an asymptomatic infection | 17 days (standard deviation 1.07)^11^ | $\log N(\text{17},\text{1.07})$ |
| $s_{i}$ | The susceptibility for different age groups | Corresponding value in Table S4 | $N_{[0,1]}(s_{i,\mu},\sigma_{s})$ |
| $\alpha_{a}^{i}$ | The ratio to be the asymptomatic infection after latent period for different age groups | Null | $Beta(2,2)$ |
| $\gamma_{i}$ | The ratio of the transmissibility of asymptomatic infection to symptomatic infection for different age groups | Null | $\mathrm{Beta}_{[0.1, 0.9]}(2,2)$ |
| *d, m* | The time that the contact starts declining and the duration of the decreasing process | Null | $\mathrm{Unif}(0, 30)$ |
| $\beta_{1},\beta_{2}$ | Correction parameter for the contact matrix | Null | $N_{[0, 2]}(1, 0.2)$ |
| $\omega_{1},\omega_{2},\omega_{3}$ | The coefficients of the B-splines in Figure S2 | Null | $N_{[0,\infty]}(0, 0.1)$ |
| $v_{1}, v_{2}$. | The parameters controlling the degree of overdispersion | Null | $Gamma(2, 1)$ |

Here $log N\left( \mu,\sigma\right)$ represents a log-normal distribution with log-mean $\text{log}\left( \mu\right)$ and log-standard deviation $\text{log}\left( \sigma\right)$. $N_{\left[ a, b \right]}\left( \mu,\sigma\right)$ represents a truncated normal distribution valued in $\left[ a,b \right]$ with mean $\mu$ and standard deviation $\sigma$. $\mathrm{IG}\left( \cdot, \cdot\right), Unif(\cdot, \cdot), Beta(\cdot, \cdot)$and $Gamma(\cdot, \cdot)$ denote the inverse Gamma distribution, Uniform distribution, Beta distribution, and Gamma distribution with their corresponding parameters, respectively. $Beta(2, 2)$, $Unif(0, 30), and Gamma(2, 1)$are flat priors for corresponding parameters, respectively. Moreover, we set $\sigma=2$*,* i.e. chose flat prior distributions for $\mathcal{L,}\mathcal{S}_{1}.$ According to the study of susceptibility for different ages^22^, we set $\sigma_{s}=0.075$ as the standard deviation of the truncated normal distributions for all age groups. The weakly informative prior distributions of ${(\gamma_{i})}_{i=1:7}$, $\beta_{1},\beta_{2}$, and $\omega_{1},\omega_{2},\omega_{3}$ are chosen to regularize the estimates of parameters for the stability of model fitting.

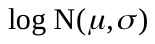

Hyper-parameters of the prior distribution of $\mathcal{S}_{2}^{i}$ are estimated from the clinical data of Zhejiang province for different age groups. To be specified, let $W_{l}^{i}$ be the observed period of symptomatic transmission for the $l^{th}$ individual in the $i^{th}$ age group, we assume an exponential mass for $W_{l}^{i}$, where $l$*=1, …,*$L_{i}$, equipped with the Jeffreys' prior for the inverse of rate parameter $\lambda_{i}$ of an exponential distribution^12^, then the posterior distribution of $\lambda_{i}$ is given as:

$$\pi(\lambda_{i}|W_{1}^{i},...,W_{L_{j}}^{i})\propto\frac{1}{\lambda_{i}}\prod_{l=1}^{L_{i}} \frac{1}{\lambda_{i}}e^{-\frac{W_{l}^{i}}{\lambda_{i}}}=IG(L_{i}, \sum_{l=1}^{L_{i}} W_{l}^{i}). (S6)$$

We use the posterior distribution of $\lambda_{i}$ as the prior distribution of $\mathcal{S}_{2}^{i}$, i.e. $a_{i}=L_{i}$ and $b_{i}=\sum_{l=1}^{L_{i}} W_{l}^{i}$. The estimated hyper-parameters are also used in the construction of the lower bound of the prior of $\mathcal{A}_{1}^{i}$. We assume that the asymptomatic infections are harder to detect than symptomatic infections after the symptom onset, i.e. $\mathcal{A}_{1}^{i}$ is longer than the mean of $\mathcal{S}_{2}^{i}$, while the upper bound of $\mathcal{A}_{1}^{i}$ is chosen as 30 since most of the asymptomatic infections will self-recover after one month^11,13–15^. For the period $\mathcal{A}_{2}$, we utilize the period from diagnosis to the first negative conversion for asymptomatic infections reported in a previous clinical study ^11,13–15^ ( a median of 17 days with a standard error of 1.07 days) to specify an informative prior distribution.

Regarding the computing algorithm, we use a random walk Metropolis-within-Gibbs algorithm^16,17^ combined with mixture transfer variance^17^ to sample the posterior distribution of the parameters of our compartmental model. A parallel computing procedure implemented by R package **snowfall**^18^ is used to efficiently generate the burn-in samples of the posterior distribution. After the convergence of MCMC, a minimum of 4000 post-burn-in samples were collected to construct the credible intervals. Subsequently, the dynamic curve of different compartments considering the posterior uncertainty could be achieved by equations (S3), (S4) and (S5) given the initial condition. These reconstructed results would be used for an illustration of the outbreak to provide Insight into the invisible dynamics of the epidemic.

#### Model assessment

We treat our structured epidemic data from January 7^th^ to February 15^th^, 2020 as a training set and use the data from February 16^th^ to February 22^nd^, 2020 as a testing set to evaluate the prediction behavior of our compartmental model. As an illustration of our model fitting, we present the fitting and prediction of the daily total new numbers of symptom onset for symptomatic infections and confirmed case for asymptomatic infections for all age in Figure S3. To measure the goodness-of-fit of our compartmental model, we use the sum of squares of residuals as the discrepancy criterion:

$$\mathrm{Dis}\left( D^{g},\Omega\right):=\sum_{t=1}^{T_{t}} \left( {D_{t}^{g} \mathcal{- D}}_{t}^{g}\left( Z\left( 0 \right),\Omega\right) \right)^{2},$$

where $D^{g}=\left( D_{1}^{g},...,D_{T_{t}}^{g} \right)$, $D_{t}^{g}=\sum_{i=1}^{7} D_{t}^{i,g}, \mathcal{D}_{t}^{g}\left( Z\left( 0 \right),\Omega\right)= \sum_{i=1}^{7} \mathcal{D}_{t}^{i,g}\left( Z\left( 0 \right),\Omega\right)$ and $T_{t}=T-7$. We use the posterior predictor assessment of model^19^ via the realized discrepancy $\mathrm{Dis}\left( D^{g},\Omega\right)$, and the Bayesian p-value for the goodness-of-fit test is calculated through the generated MCMC samples.

Our results show a good fit to the daily new numbers of symptom onset cases (Figure S3 (A)) and daily new confirmed asymptomatic infections (Figure S3 (B)). The Bayesian p-values of the goodness-of-fit test are 0.451 and 0.180, respectively, implying that there is no evidence against the fitted curves. Specially, the observed numbers in the prediction period (Figure S3) are both covered by the credible bands, which indicate the reliability of the proposed compartmental model and simulation results.

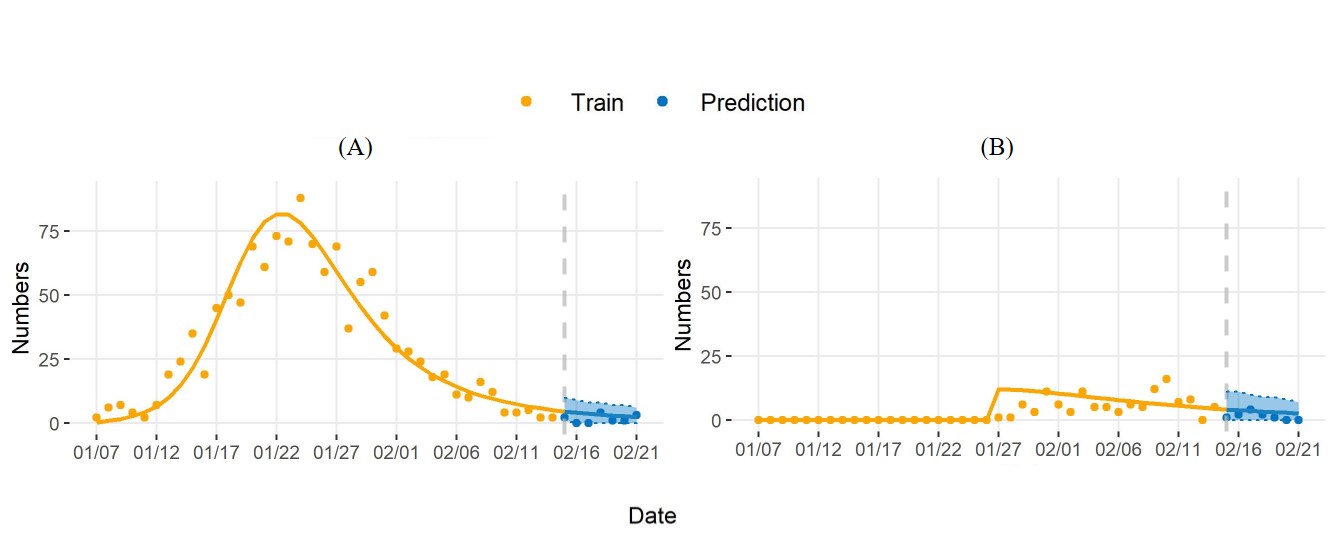

Figure S3. The daily total new numbers of symptom onset for symptomatic infections (A) and confirmed case for asymptomatic infections (B) for all age groups from January 7^th^ to February 21^st^, 2020**.** The fitting curve form January 7^th^ to February 14^th^, 2020 and the predicted numbers with 95% credible band form February 15^th^ to February 21^st^, 2020 are presented, where the fitting and predicted numbers are calculated by the means of posterior distributions.

#### Supporting results

Age-varying transmissibility

The estimated age-varying transmissibility of asymptomatic and symptomatic infections of Figure 2 are given in Table S6 and Figure S7.

Table S6. The estimated transmissibility and the ratios of the medians of their posterior distributions ($\frac{T_{i,a}}{T_{i,s}}$) in different age groups

|  | The transmissibility of symptomatic infection ($T_{i,s}$) | The transmissibility of asymptomatic infection ($T_{i,a}$) | Ratio of transmissibility $T_{i,a}/T_{i,s}$(%) |
| --- | --- | --- | --- |
| 0-10 | 0.043 (0.014, 0.123) | 0.016 (0.003, 0.071) | 35.70 |
| 10-20 | 0.058 (0.029, 0.116) | 0.021 (0.006, 0.062) | 35.73 |
| 20-30 | 0.078 (0.050, 0.124) | 0.024 (0.008, 0.065) | 30.35 |
| 30-40 | 0.107 (0.077, 0.149) | 0.026 (0.011, 0.077) | 24.42 |
| 40-50 | 0.144 (0.107, 0.186) | 0.034 (0.015, 0.093) | 23.38 |
| 50-60 | 0.189 (0.142, 0.241) | 0.055 (0.020, 0.152) | 29.04 |
| 60+ | 0.243 (0.182, 0.309) | 0.132 (0.036, 0.224) | 54.33 |

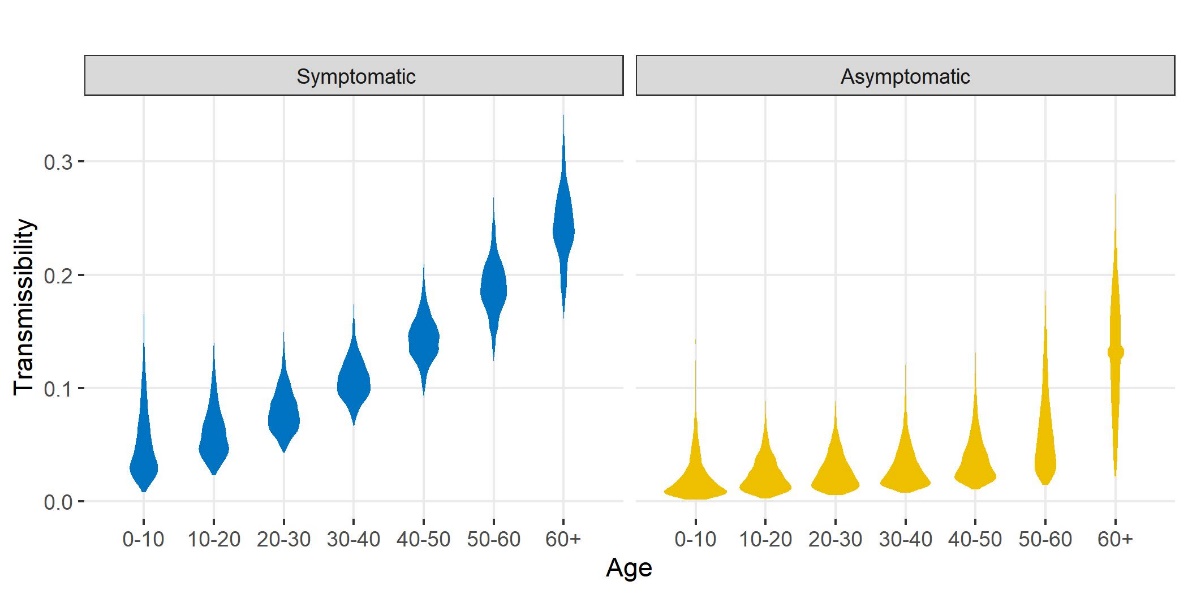

*Figure S4. The eye-plot of the posterior distributions of transmissibility for seven age groups.*

Typically, the estimated transmissibility was identified based on the reported susceptibility, which implied elder persons were more susceptible than the younger people^20^. This relative measure would be helpful for us to the evaluate the true intensity of shedding virus for infectious individuals.

Unconfirmed numbers

We present the observed numbers of symptomatic cases, asymptomatic cases and the estimated unconfirmed numbers of asymptomatic infections for different age groups in Table S7, which are used for the calculation of Figure 3.

Table S7. The infected individuals until February 22^nd^, 2020 for different age groups

|  | Observed numbers of symptomatic cases | Observed numbers of asymptomatic cases | Estimated unconfirmed number of asymptomatic infections |
| --- | --- | --- | --- |
| 0-10 | 15 | 10 | 12.67 (7.33, 20.44) |
| 10-20 | 24 | 10 | 22.66 (11.67, 38.68) |
| 20-30 | 123 | 14 | 90.39 (39.12, 165.37) |
| 30-40 | 259 | 22 | 75.21 (41.83, 133.10) |
| 40-50 | 272 | 23 | 63.21 (34.72, 108.29) |
| 50-60 | 294 | 23 | 44.19 (25.55, 70.60) |
| 60+ | 231 | 22 | 40.61 (24.05, 68.13) |
| Total | 1218 | 124 | 354.30 (239.11, 519.30) |

Number of newly infected caused by different transmissions

The estimated results related to Figure 4 and 5 are given in Table S8 and S9, which are estimated from equations (S4) and (S5).

Table S8. The newly infected numbers caused by symptomatic $E_{s}\left( t \right)$ and asymptomatic transmission $E_{a}\left( t \right)$during different time period

|  | Infected numbers caused by symptomatic transmission | Infected numbers caused by asymptomatic transmission | Proportion of the asymptomatic transmission (%) |
| --- | --- | --- | --- |
| 01/07-01/11 | 74.18 (59.53, 92.56) | 6.25 (3.24, 10.85) | 7.77 |
| 01/12-01/16 | 415.47 (321.21, 527.94) | 57.85 (29.21, 100.43) | 12.22 |
| 01/17-01/21 | 739.91 (656.73, 844.47) | 110.71 (61.06, 179.17) | 13.02 |
| 01/22-01/26 | 276.81 (195.25, 370.57) | 46.11 (25.41, 75.09) | 14.28 |
| 01/27-01/31 | 38.01 (20.84, 64.27) | 7.26 (3.57, 13.05) | 16.03 |

Table S9. The infected numbers caused by different age groups until February 1^st^, 2020

|  | Contribution rate for symptomatic transmission (%) | Contribution rate for asymptomatic transmission (%) |
| --- | --- | --- |
| 0-10 | 0.09 (0.03, 0.32) | 0.49 (0.09, 2.59) |
| 10-20 | 0.41 (0.19, 0.90) | 1.99 (0.42, 7.84) |
| 20-30 | 3.77 (2.55, 5.70) | 9.44 (2.62, 27.74) |
| 30-40 | 18.53 (15.19, 22.75) | 16.52 (5.84, 40.98) |
| 40-50 | 21.65 (18.70, 24.92) | 14.74 (5.55, 37.32) |
| 50-60 | 28.63 (25.19, 32.16) | 16.49 (5.67, 40.69) |
| 60+ | 26.55 (21.65, 31.36) | 31.73 (10.19, 56.10) |

Simulation studies

Simulation studies are conducted to further investigate the dynamic changes in transmission burden over time during a prolonged epidemic (Figure S5). When the duration of the decreasing process of contact function (represented by “$m$” in Figure S1) was prolonged by two weeks and each individual’s daily contact number increased by one person during the outbreak period, we found a slower decreasing trend in daily new cases. Furthermore, the proportion of asymptomatic infections contributing to the spread of the virus increased during the early outbreak and period in which daily new cases are decreasing (Figure S5, scenario 1), which implies that although the early outbreak was mostly contributed to by symptomatic infections, asymptomatic transmission would increase rapidly thereafter and continue to rise along with the constant outbreak of COVID-19.

We further proposed two more simulations (Figure S5, scenario 2 and scenario 3). In order to promote the generalizability of our model to better guide interventions in other regions, we let the time needed for tracing the symptomatic cases shrink 50% based on scenario 1 above (scenario 2), and then let one jurisdiction have limited ability to trace contacts, thereby being generally unable to find asymptomatic cases based on scenario 2 (scenario 3). In scenario 2, quicker isolation effectively suppressed the spread of COVID-19, while the burden of asymptomatic transmission increased. In scenario 3, asymptomatic transmission dominated the total transmission when the number of daily new infected cases was still considerable.

Overall, we found either symptomatic or asymptomatic transmission could dominate the outbreak (Figure S8). If strong controlling measures are solely implemented on symptomatic cases, the unrestricted asymptomatic transmission will likely prolong the needed time of interventions and potentially delay reopening.

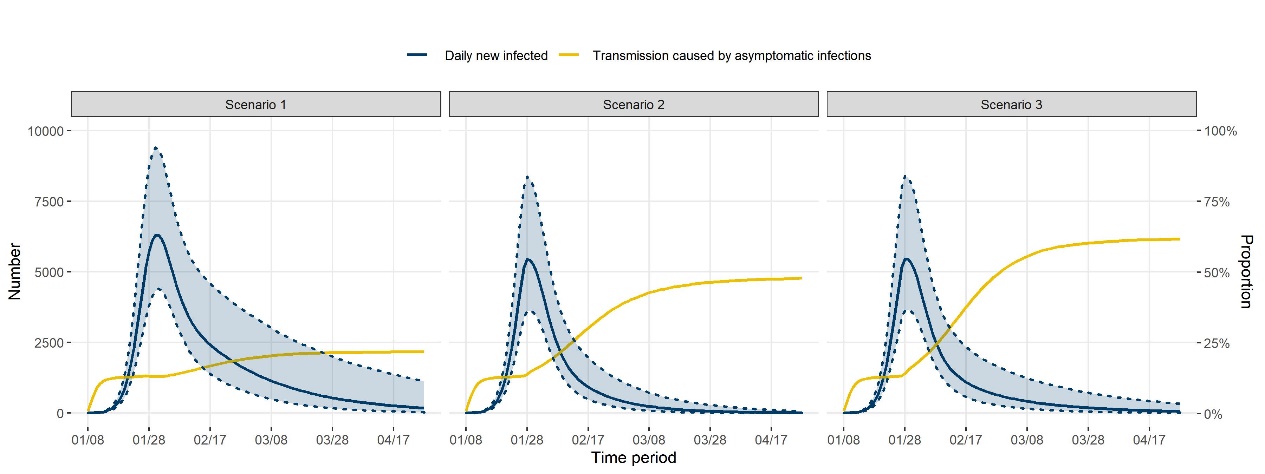

*Figure S5. The simulated daily new infected numbers and their proportion of new infected cases caused by asymptomatic transmission (from January 7^th^ to April 27^th^, 2020), where* ***scenario 1*** *assumed the estimated duration of the decreasing process of contact matrix was prolonged by two weeks and average contact numbers per day for each age group was increased by 1 during the lockdown period.* ***Scenario 2*** *assumed the duration between symptom onset and confirmation by nucleic testing (represented by* $\mathcal{S}_{2}^{i}$*) for symptomatic infections was shortened 50% after January 28^th^ based on scenario 1.* ***Scenario 3*** *assumed the asymptomatic infections were not controlled (i.e.* $1/{\mathcal{A}_{1}^{i}}=0$*) based on scenario 2. The Y-axis on the left denoted the numbers of daily new infected and the Y-axis on the right represented the proportion of the transmission among daily new infected caused by asymptomatic infections. The 95% credible intervals for simulated numbers are shown for each scenario.*

To visualize the differences of the contribution rate in Figure 5 among different age groups, we proposed seven scenarios of simulations to evaluate the potential outcome if one age group was immune to the COVID-19 by vaccine (Figure S6). If the vaccine could significantly decrease the susceptibility and thereby transmissibility due to not being infected, the numbers of all infections decreased 2.09%, 3.95%, 26.27%, 59.89%, 66.33%, 79.35%, 55.44% in each age group, respectively.

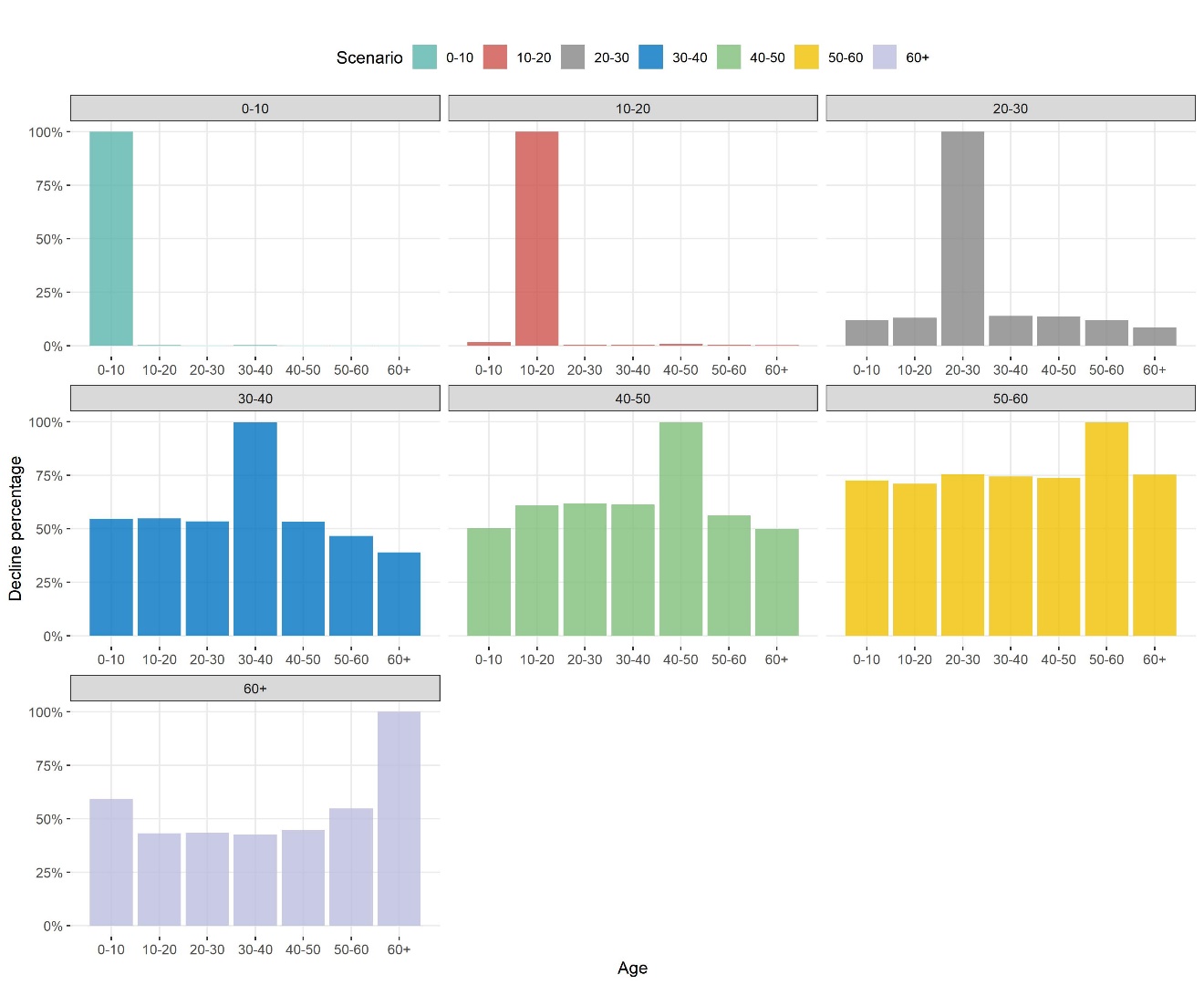

*Figure S6. The simulated decline percentages of all infected individuals in different age groups (the population except the susceptible individuals) until February 22^nd^, 2020 compared with the original estimates in a different scenario. Each scenario assumed one of the age groups was immune to COVID-19, i.e. not susceptible and transmissible to COVID-19, respectively.*

1. The mean of the susceptibility of “60+” is calculated as the average of the mean for age groups “60-69” and “70+” in that study.
